# Optimising the Usability of AI Driven Augmented Reality Displays of Critical Structures During Surgery – An International Study of Surgeon-Computer Interaction

**DOI:** 10.64898/2026.06.02.26354758

**Authors:** Roxana Ramirez Herrera, Danyal Z. Khan, Anjana Wijekoon, Sophia Bano, Matthew J. Clarkson, Hani J. Marcus, Ann Blandford, CARES Evaluation Group

## Abstract

In many endoscopic surgical procedures, the surgical team must identify and remove pathological tissue while avoiding critical structures such as arteries and nerves. Augmented reality (AR) offers potential support by overlaying visual information about the location of pathology and critical structures directly onto the operative field, enhancing spatial awareness and surgical navigation. However, limited research has evaluated how best to design and present AR overlays in ways that align with surgical workflow and perception. This study investigates surgeons’ preferences across three key AR overlay dimensions: Design (how anatomy is visualised: outlines, heatmaps, masks, or centroids), Trigger (how and when overlays are activated: always visible, activated by the user, or triggered by instrument position), and Placement (where the overlay appears: above or below the surgical instrument). We take endoscopic pituitary adenoma surgery as a high-risk exemplar. Using a web-based prototype, 38 neurosurgeons ranked options and provided qualitative feedback. Surgeons preferred outline designs for clarity, user-activated triggers for control of information flow and distraction minimisation, and below-instrument placement for better spatial awareness. Preferences were consistent across experience levels and emphasised the importance of balancing visual saliency with cognitive load, to facilitate surgical navigation without distraction or disruption. These findings inform AR interface design, but require evaluation for impact on surgical performance and safety in further physical simulation and clinical studies.

- International evaluation of surgeon AR preferences in high-risk neurosurgery.
- Surgeons prefer outline AR design for clarity and minimal field obstruction.
- Manual triggers are preferred to maintain surgeon autonomy and reduce distraction.
- Design must balance visual saliency with the surgeon’s cognitive load.

## 1. Introduction

Augmented Reality (AR) technologies are increasingly being explored to support intraoperative navigation and decision-making in surgery. AR can enhance spatial awareness and depth perception by overlaying anatomical information directly onto the surgical view, particularly in procedures with limited visual access. While AR has been studied in other surgical domains, neurosurgery has kept its development at the forefront with applications ranging from spinal surgery and vascular navigation to neuronavigation and skull base approaches [1]. These procedures often involve navigating through narrow corridors surrounded by critical structures, where margins for error are extremely small. In this context, the design and timing of AR overlays become critical: systems must provide intuitive and context-relevant information without obstructing the field of view or increasing cognitive load [2, 3, 4]. These challenges are particularly relevant in endoscopic neurosurgery, where the surgical field is typically visualised on a 2D screen and the space for additional instruments (e.g., image-guidance or Doppler probes) is limited [2].

A key example of such a procedure is the endoscopic TransSphenoidal Approach (eTSA), which provides access to the pituitary gland via the nasal cavity and serves as a valuable case study for examining AR design. Pituitary adenomas are among the most common primary brain tumours, with a prevalence of up to 20% [5]. Although benign, these tumours are often locally invasive and can encase or distort surrounding structures such as the carotid arteries, optic nerves, and pituitary gland. The eTSA is the standard surgical method for tumour removal, but it carries significant risk: incomplete resection (up to 70%), vascular injuries (1–2%), and new hormonal deficits (up to 50%) [6, 7]. These complications can have substantial consequences, including increased morbidity, longer hospital stays, and the need for further interventions. Navigation tools such as image guidance and Doppler ultrasound are commonly used in the sellotomy phase of this operation where these structures are most at risk, but these have limitations, including intraoperative tissue shifts and workflow disruption [8]. Given these constraints, pituitary surgery offers a compelling case study for evaluating how AR may support safer, more efficient intraoperative navigation.

In response to these challenges, our group has developed an Artificial Intelligence (AI)-based AR system that overlays anatomical landmarks onto the endoscopic video feed in real time. In preclinical simulation studies, we have shown that this system can help surgeons recognise key structures with improved clarity during pituitary surgery [9, 10, 11]. The AI-based AR system is designed to operate without introducing additional hardware into the surgical field, and it integrates seamlessly into the operative workflow. However, the optimal way to present this visual information via AR - what to show, when to show it, and where to place it - remains an open question. Because neurosurgery is a high-stakes, dynamic environment, interface design decisions must account for cognitive demands, spatial reasoning, and the potential for attentional overload.

In this paper, we investigate neurosurgeons’ preferences across three key dimensions of the AR overlay interface for eTSA: Design (how visual information is presented, such as heatmap or outline), Trigger (how and when the overlay is activated), and Placement (where the overlay appears in relation to the surgical instrument). We focus specifically on the sellotomy phase—when the sella turcica is opened to access the pituitary region—during which accurate identification of anatomical landmarks is critical and AR support is most impactful [12]. The dimensions and their corresponding variants were identified through discussions with clinicians and computer scientists involved in the project. Using a web-based prototype, we presented each variant to the participants and asked them to rank their preferences. We collected both quantitative and qualitative data from neurosurgeons with varying levels of experience and from different countries. While pituitary surgery serves as the motivating case study, the interaction challenges identified—visual saliency, user control, timing of information, and cognitive load are broadly applicable to safety critical augmented reality and AI-supported decision-making environments, and inform the design of a context-aware, user-centred AR interfaces for surgery.

## 2. Related Work

Overlays of critical anatomical landmarks have become the standard implementation of AR in eTSA, achieved through various methods of segmentation and registration [13, 14]. While the literature frequently reports that AR enhances confidence in decision-making during surgery and shows high levels of acceptability, there has been limited evaluation of *how* information is presented. Challenges around how such visualisations are presented are not unique to eTSA. Across a range of surgical domains—including laparoscopy, spine surgery, and neuronavigation—surgeons must interpret augmented content within constrained visual fields while avoiding critical structures [1]. This highlights broader Human-Computer Interaction (HCI) concerns about how AR systems can support decision-making without introducing unnecessary complexity. Rather than focusing solely on technical accuracy or tracking performance, there is a growing need to address how information is surfaced to users and how it aligns with their cognitive, perceptual, and ergonomic needs [15].

One major concern across surgical AR is that poorly designed overlays can lead to visual clutter and distract from the operative field. This risk has been discussed in both conceptual and empirical studies, which show that cluttered overlays can increase cognitive load and even lead to inattention blindness [16, 17, 18]. As AI-enabled AR tools become more advanced, incorporating HCI principles during early design stages is essential to ensure that these systems address real user needs, rather than focusing solely on technical feasibility [19].

Several studies have begun to explore how surgeons engage with AR interfaces and which visualisation styles are most effective. For instance, depth perception has emerged as a central concern in AR visualisation. Heiliger et al. (2023)[20] compared five vascular visualisation techniques in laparoscopic AR and found that 55% of participants preferred high-opacity renderings with single-colour Fresnel highlights, which appeared to enhance spatial clarity. Similarly, Sielhorst et al. [21] assessed surgeons’ ability to perceive depth in medical AR environments and emphasised the limitations of systems lacking adequate 3D cues. Nguyen et al. [22] evaluated visualisation preferences using the Microsoft HoloLens and found that experienced surgeons preferred grey-scale overlays aligned with conventional imaging, whereas less experienced users favoured more colourful schemes. The same study also revealed a 95% preference for high-opacity overlays of hard tissue, while soft tissue overlays were less favoured and sometimes actively avoided.

In addition to visualisation style, interaction methods have also been evaluated for their intuitiveness. Sánchez-Margallo et al. [23] found that a MYO armband combined with voice commands was rated the most intuitive and accurate, whereas Kinect-based gesture controls were considered physically demanding and prone to unintended inputs. Usability and ergonomic comfort have similarly been assessed through subjective ratings and observational data. Iqbal et al. [24], for instance, reported positive feedback on usability and minimal physical discomfort with optical see-through head-mounted displays in robot-assisted orthopaedic surgery. However, Nguyen et al. [22] noted that current AR headsets were not well-suited for prolonged use due to issues such as eye fatigue and difficulty adapting to changing lighting conditions in the OR. Design strategies specific to eTSA have also been explored. Higa et al. [25] investigated how partial anatomical visualisations could enhance trust and reduce cognitive load during endoscopic pituitary surgery. Their findings suggest that more information is not always better—selective overlays may help optimise clarity and decision-making.

The concept of context-aware AR was introduced to address the issue of visual clutter. In this paradigm, computational strategies are employed to identify specific surgical phases or instrument use [26, 27]. This approach enables the system to display only the information relevant to the current action, rather than all the information for the entire procedure [26, 27]. Although predictive systems could eventually determine the information that surgeons expect at a certain step or phase of surgery, there will still be instances where surgeons need to access information of their choosing. Additionally, the surgeon’s skill level may dictate how much additional information is necessary.

However, how system capabilities to filter information and visualisations can better serve surgeons’ information needs remains an area that requires further exploration. Furthermore, AR approaches to support surgery have mostly provided an all-or-nothing solution, where users can only toggle AR overlays on or off, limiting control over information visualisation throughout the procedure after the initial setup [13, 28]. This highlights a gap in research on how different interaction modalities could leverage AR usage, increase confidence in decision-making, and improve system usability. To be feasible in real-time, such systems must remain efficient. High-fidelity 3D overlays can place bandwidth and processing power demands if not optimised, and have not been accurately and automatically registered to live 2D surgical videos [29, 30]. As a result, minimalistic design principles are crucial to ensure clarity and usability. The interface should present only essential information, reducing visual clutter to avoid overwhelming the surgeon [19]. As noted above, preliminary studies by Higa et al. [25] suggest that surgeons prefer partial visualisations of anatomical landmarks to enhance trust and reduce cognitive load. This preference implies that a carefully curated selection of information, providing just enough detail to navigate complex tasks without clutter, could be key to optimising the utility of AR during surgery. Therefore, this paper explores how surgeons with varying levels of experience perceive different design options for an AI-driven AR interface intended to support critical structure recognition. By using endoscopic pituitary adenoma resection as a case study, we aim to contribute to ongoing HCI efforts that prioritise usability, clarity, and adaptability in surgical AR systems.

## 3. Methods

To explore surgeon perspectives and preferences for AR interface design, we employed a cross-sectional study design, via an interactive online mixed methods survey. As pituitary surgery is concentrated on a relatively small number of specialists per neurosurgical centre, an online format was chosen to facilitate the participation of surgeons from various centres.

### 3.1. Participants

The survey was disseminated via multiple society mailing lists – including the Society of British Neurological Surgeons (SBNS), British Neurosurgical Trainees’ Association (BNTA), British Neuroendoscopy Society (BNES) and social media (X, LinkedIn) on channels of relevance for neurosurgeons. It was also disseminated through direct word of mouth and mailing lists within University College London (UCL). The eligibility criteria to participate were: neurosurgeons with at least one year of experience performing the eTSA and able to communicate confidently in English. Participants were offered collaborative group authorship to recognise their contribution to the work. The survey remained open for data collection for three months from 5 October 2024 to 5 January 2025.

A total of 38 neurosurgeons fulfilling the entry criteria participated in the survey – see Figure 1. Attending or consultant-level professionals made up 53% of the participants, while 16% were senior fellows who had completed their neurosurgical training, and the remaining 31% were still in training. The median years of experience in pituitary surgery was 4 years with interquartile range (IQR) 2 – 9.5 years. Most participants were from the United Kingdom (UK) (68%, 26/38), reflecting our primary recruitment channels. Other participants were 13% (5) from Belgium, and 3% (1) each from Egypt, Germany, Italy, Saudi Arabia, Spain, Switzerland, and Turkey.

**Figure 1:**
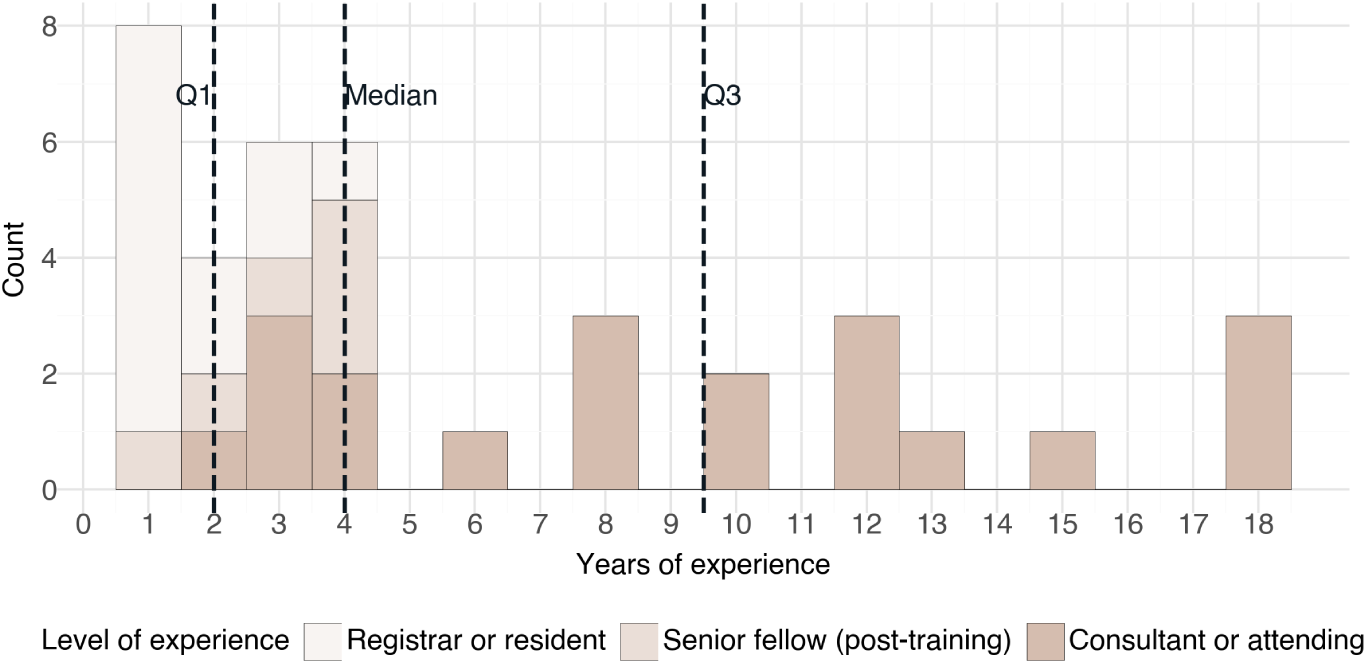
Histogram of participants’ years of experience and level of experience; Median was 4 years of experience with 2–9.5 IQR.

### 3.2. Hypotheses

We investigate the following hypotheses:

- H1: There exists a statistically significant preference for an AR display type – Outline, Mask, Heatmap, or Centroid – over the others.
- H2: There exists a statistically significant preference for an AR trigger type – user-enabled (sella & parasellar structures), instrument-triggered (parasellar) & sella user-enabled, instrument-triggered (parasellar) & sella always on or always-on (sella & parasellar structures) – over the others.
- H3: There exists a statistically significant preference for one AR placement – over the instruments or below the instruments – over the other.

### 3.3. Survey Design

The survey was co-designed by an interdisciplinary team of surgeons, human-computer interaction scientists and computer scientists, with a low-fidelity web-based AR prototype developed in Framer ^3^ and embedded within a Qualtrics survey (for full survey questionnaire, see Appendix A) with standard security measures [31]. The clinical context selected was the “sellar phase” of the eTSA, which is the section of the surgery during which surgeons remove the tumour whilst protecting surrounding critical structures from injury. Endoscopic images from this phase were selected from an open-access video database of eTSA [32]. As these surgeries are performed using a 2D endoscope, the resulting images are 2D stills and were annotated for critical anatomical structure outlines by expert surgeons. The selection and annotation of these structures were based on prior consensus and surgical computer vision studies [12, 9, 10, 11]. The survey format was as follows. After being given information about the study background and aims, and consenting to participate, demographics (country of practice) and clinical experience (grade, years of pituitary surgery experience) were gathered. The survey was then presented in three sections to evaluate three main design domains: 1) overlay design, 2) overlay trigger, and 3) overlay placement.

#### 3.3.1. Overlay Design

In this section, participants were presented with four different AR visualisations of both sella (the region where the tumour is centred) and the surrounding critical “parasellar structures” (carotid arteries, optical protuberances and clival recess). The visualisation options – Centroids, Safety Outline, Masks, Heatmaps - described in Figure 2 resulted from an iterative design process within the interdisciplinary group reviewing precedence within the surgical anatomical structure segmentation literature [4, 10, 11], filtered by what was technically feasible to implement in real-time at present (e.g., 3D overlays registered to 2D endoscopic images were excluded) [25]. The participants were presented with the 2D visualisation options overlayed on still surgical images (see Figure 2), with the ability to toggle between the above options for the “sella” and the surrounding “parasellar” critical structures separately. After toggling through the options, they were asked to rank their visualisation preferences from most to least preferred (1 to 4) and provide written feedback explaining their choices for most and least liked visualisations.

**Figure 2:**
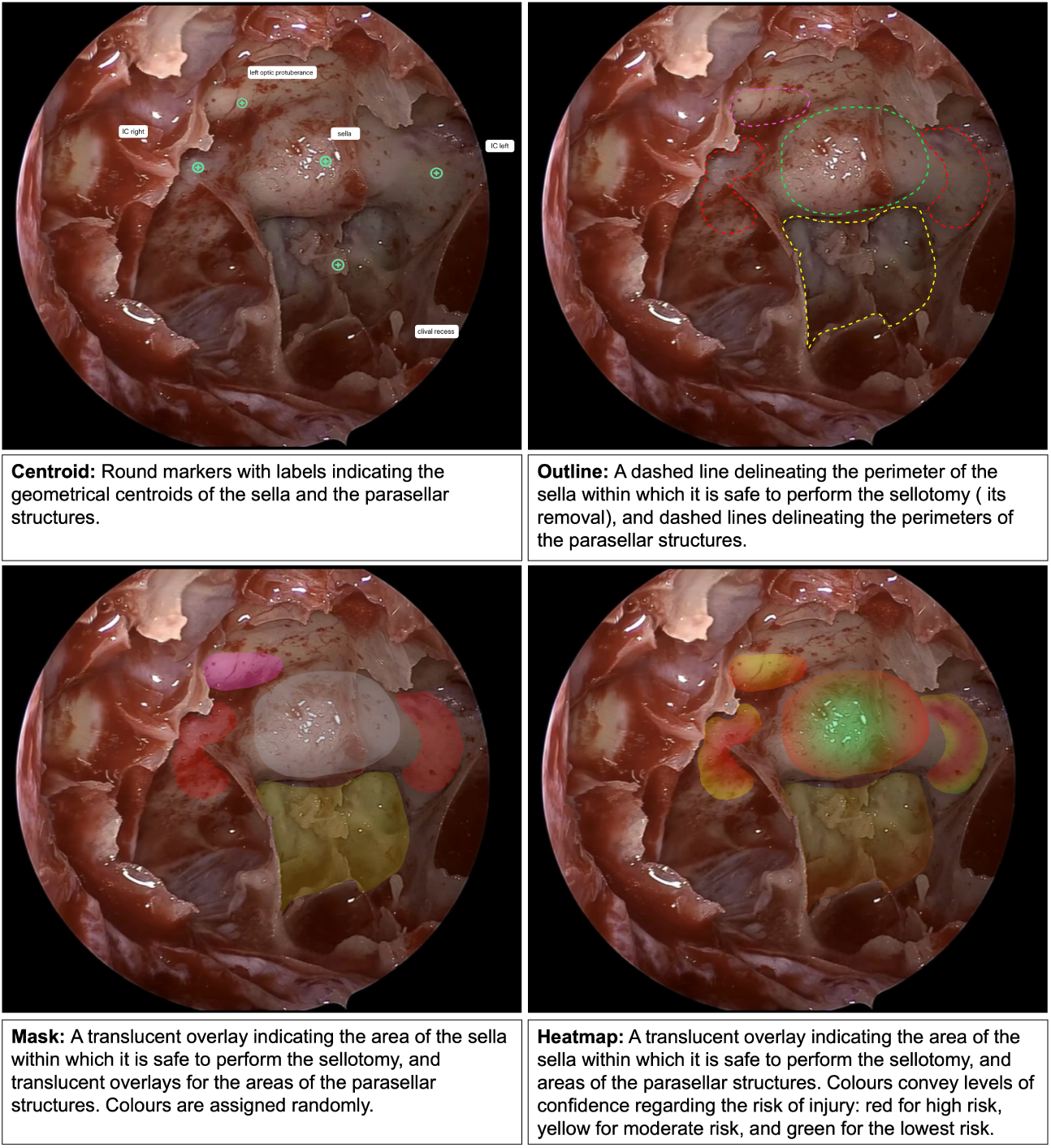
Overlay Designs; Descriptive text for each design appeared as a pop-up when selected by participants.

#### 3.3.2. Overlay Trigger

In this section, participants were presented with four options for triggering visualisations, which control the appearance and disappearance of the supporting AR, as described in Figure 3. The trigger options provide different levels of agency to the user; they were derived from the literature and refined through internal interdisciplinary consensus, and consisted of: 1) always on (sella & parasellar structures); 2) user-enabled (sella & parasellar structures); and instrument-triggered for parasellar structures (overlay appears only when the instrument comes close to that critical structure) 3) with sella always on and 4) with the sella user-enabled. For the assessment of these triggers, participants were asked to perform a simulated sellotomy using the mouse (which was rendered on the screen as a virtual diamond drill, see 3) to paint the area in which they would drill on that image to access the tumour safely. They tested these visualisation triggers with their first choice of design for sella and parasellar from the previous section. They performed this four times, one run per trigger option, and then ranked the options from most to least preferred (1 to 4) with free text justification of their least and most preferred choices. Afterwards, they were asked to rank on a Likert scale from 1 to 5 (strongly agree to strongly disagree) to evaluate each visualisation trigger on how helpful and distracting they found it.

**Figure 3:**
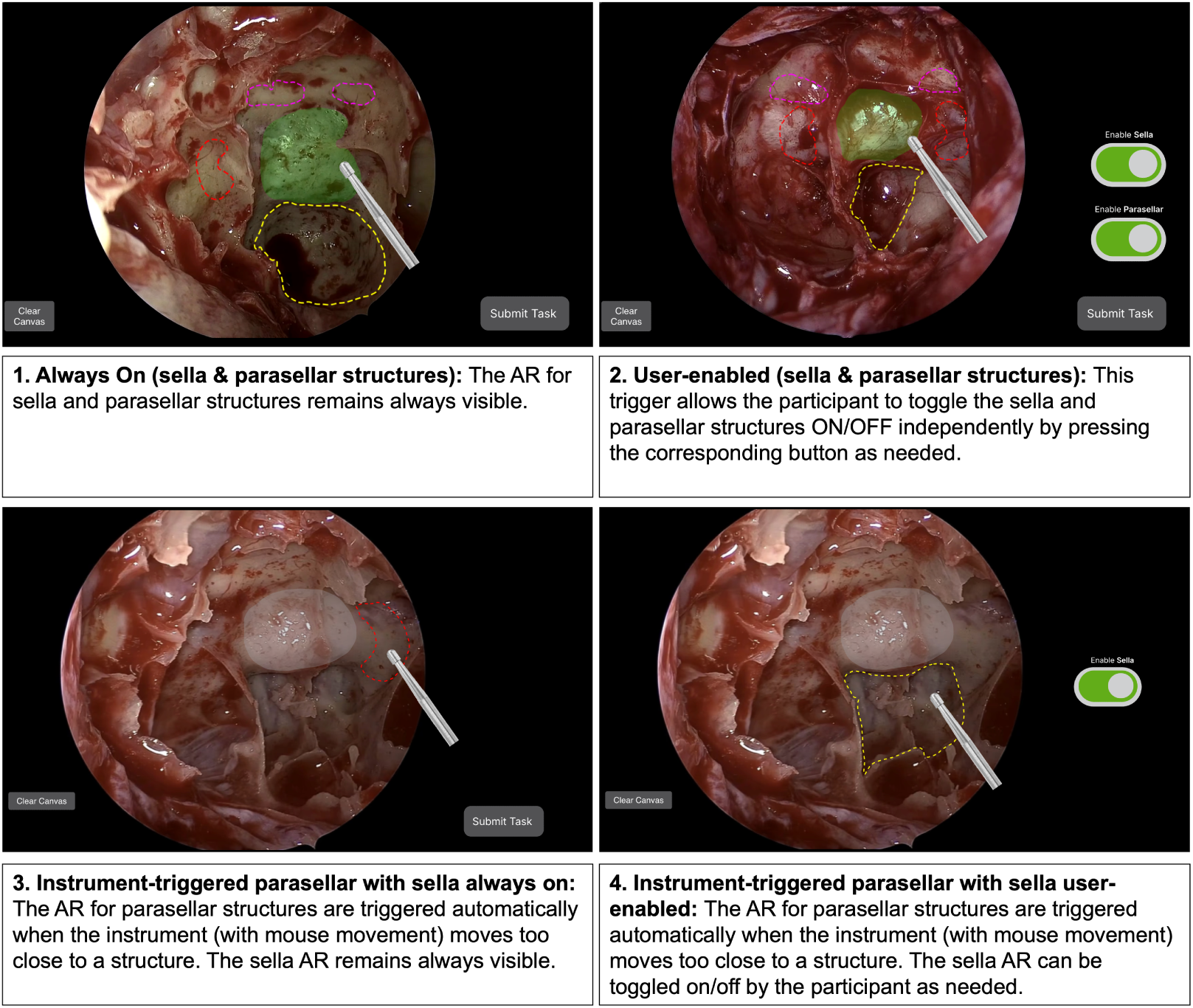
Overlay Triggers; The examples selected represent a scenario where the participant selected Mask and Outline as their top-ranked overlay designs in the previous section.

#### 3.3.3. Overlay placement with respect to surgical instrument

Overlay placement refers to whether the visualisation is presented to the user over the instrument (as an overlay) or below the instrument (as an underlay) – see Figure 4. In this scenario, the mouse cursor was rendered again as a virtual drill, which participants could move around the endoscopic image while toggling whether the AR visualisation was placed over or below the instrument. For this task, participants were not shown their preferred visualisation as in the previous task. Instead, they were presented with a combination of Outline and Mask to enhance instrument placement visibility. This combination provided sufficient visual coverage, whereas designs such as centroids would not have provided a meaningful comparison. The combination also ensures the difference in layering is more salient than the Outline or Mask alone. They were asked their preference between the two options, and to justify with free text. Finally, the participants were invited to give wider feedback on the designs and whether they felt any additional options should be considered in an open-ended question.

**Figure 4:**
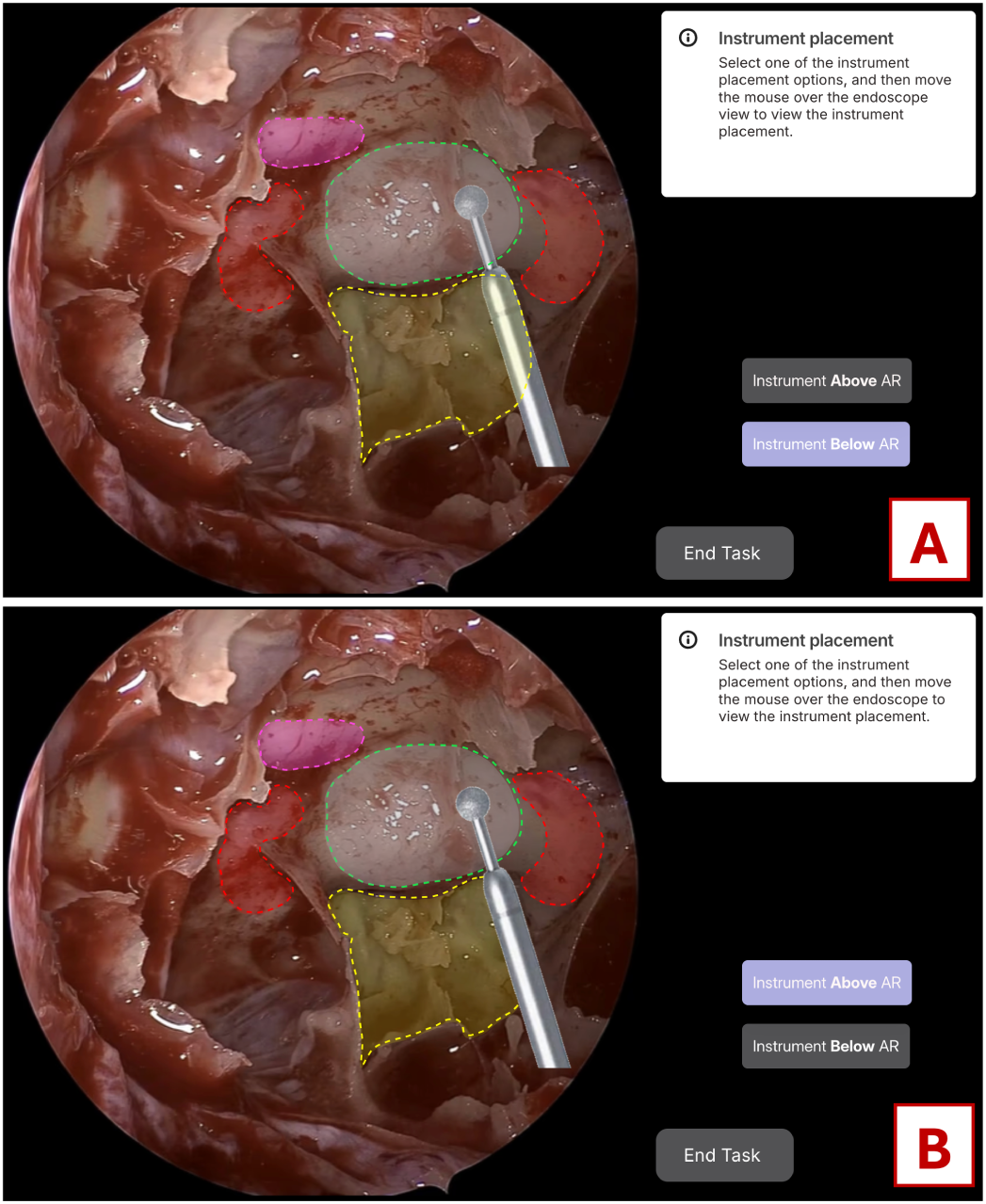
Overlay placements: (A) overlay above the instrument; and (B) overlay below the instrument. Overlay designs were chosen as a combination of Outline and Mask to enhance instrument placement visibility during this task.

### 3.4. Data Analysis

Quantitative data (rankings and Likert-scale questions) were summarised using descriptive statistics and further analysed using inferential statistics to determine statistically significant preferences in overlay design, triggers and placement.

Considering the distribution of participants’ experience levels, first, we ensured that all participants were a sample of a single population. For this, we identified two sub-groups of residents/fellows vs consultants and applied the following tests: a) the Friedman test to check within-group ranking differences; b) Mann-Whitney U test to compare the rank distributions between groups; and c) Ordinal Logistic Regression (OLR) to test the interaction effect between groups. A Wilcoxon Signed-Rank Test was applied to test each hypothesis. Friedman, Mann-Whitney U and Wilcoxon Signed-Rank tests were implemented using scipy Python library ^4^ and ordinal logistic regression was implemented using statmodels Python library ^5^.

For the qualitative data, a thematic analysis of open-ended question responses using MAXQDA 24 ^6^ was conducted. Responses were categorised using deductive coding based on overlay design, trigger, placement, and whether participants expressed a preference or dislike. Additionally, inductive coding was used to identify subthemes reflecting participant perceptions. Finally, themes were grouped into broader insights to better understand user preferences and concerns. To support theme development and guide the structure of the findings, code intersections with overlay design, trigger, and placement were explored using the code relations browser and matrix tools in MAXQDA 24. These visualisations of code intersection and frequency helped identify prominent patterns across design elements and informed the grouping and relative emphasis of specific subthemes. The coding was carried out by one of the authors; as a result, inter-coder agreement measures were not applicable.

### 3.5. Ethics Considerations

This study was approved by the University College London Interaction Centre (UCLIC) Ethics Board with study registration number UCLIC-2024_009. All participants were fully informed about the study, participated voluntarily and provided written consent through the online survey. All data was stored and managed per current UK GDPR standards [33].

## 4. Results

The results are presented in two parts for each of the three design categories evaluated in the survey: overlay design, overlay triggers, and overlay placement. For each category, the quantitative findings, including descriptive statistics and inferential testing, are presented first, followed by the qualitative thematic analysis. Within the qualitative findings, participants are referred to as P1 through P38 where, for example, P1 corresponds to Participant 1, P2 to Participant 2, etc.

### 4.1. Overlay Design

Regarding sella overlay designs, Outline was ranked highest on average and most frequently ranked first, whilst Centroid was ranked the lowest on average and most frequently ranked last (Table 1). Subgroup analysis revealed no statistically significant differences in item rankings between Residents/Fellows vs Consultants (Friedman test: p=0.999 for each group; Mann-Whitney U test: p=1.0). OLR further supported this and showed no significant effect of group on ranking with p = 0.969. Post-hoc analysis via Wilcoxon Signed Rank testing showed that preference for Outline was statistically significant over Mask (p=0.001), Heatmap (p=0.024) and Centroid (p<0.001). Mask was preferred over Centroid (p<0.01) but there was no significant difference in preference over Heatmap (p=0.491). Centroid was the least preferred with statistical significance over all other designs.

**Table 1:** Analysis of Sella overlay design rankings; Rows ordered by quartile statistics. Outline is the most preferred design both from ranking count and median. The preference for outline over Mask is statistically significant (highlighted in bold text). Centroid is the least preferred over Heatmap with statistical significance (underlined text).

| Design | Count |  |  |  | Quartile statistics |  |  | Wilcoxon test - p value |  |  |  |
| --- | --- | --- | --- | --- | --- | --- | --- | --- | --- | --- | --- |
|  | Rank 1 | Rank 2 | Rank 3 | Rank 4 | Median | Q1 | Q3 | Outline | Mask | Heatmap | Centroid |
| Outline | 20 | 8 | 10 | 0 | 1 | 1 | 2.75 | – | **0.001** | 0.024 | 0.000 |
| Mask | 5 | 16 | 16 | 1 | 2 | 2 | 3 | **0.001** | – | 0.491 | 0.000 |
| Heatmap | 13 | 6 | 6 | 13 | 2.5 | 1 | 4 | 0.024 | 0.491 | – | <u>0.005</u> |
| Centroid | 0 | 8 | 6 | 24 | 4 | 3 | 4 | 0.000 | 0.000 | <u>0.005</u> | – |

For parasellar overlay designs, Outline achieved the highest mean rank and was most frequently ranked first, while Centroid had the lowest mean rank and was most frequently ranked last (Table 2). Subgroup analysis showed no statistically significant difference between subgroups with Friedman test p=0.999 for each group; Mann-Whitney U test: p=1.0 and OLR effect of group on ranking p=0.969. Comparative analysis using Wilcoxon Signed Rank testing showed that Outline was preferred over Centroid (p<0.01) and heatmap (p<0.01) but without a statistically significant difference over Mask (p=0.24). Preference for Mask over Centroid (p<0.001) and Heatmaps (p=0.009) were statistically significant. The least preferred design Centroid was not significantly different from Heatmap design (p=0.184).

**Table 2:** Analysis of Parasellar overlay design rankings; Rows ordered by quartile statistics. Outline is the most preferred design from the rank count while Mask achieves a similar median. The preference for Outline over Mask was not statistically significant (bold text). The ranking of Centroid as the least preferred design is also not statistically significant over Heatmap (underlined).

| Design | Count |  |  |  | Quartile statistics |  |  | Wilcoxon test - p value |  |  |  |
| --- | --- | --- | --- | --- | --- | --- | --- | --- | --- | --- | --- |
|  | Rank 1 | Rank 2 | Rank 3 | Rank 4 | Median | Q1 | Q3 | Outline | Mask | Heatmap | Centroid |
| Outline | 18 | 9 | 10 | 1 | 2 | 1 | 3 | – | **0.243** | 0.005 | 0.000 |
| Mask | 10 | 16 | 10 | 2 | 2 | 1.25 | 3 | **0.243** | – | 0.009 | 0.000 |
| Heatmap | 8 | 5 | 12 | 13 | 3 | 2 | 4 | 0.005 | 0.009 | – | <u>0.184</u> |
| Centroid | 2 | 8 | 6 | 22 | 4 | 2.25 | 4 | 0.000 | 0.000 | <u>0.184</u> | – |

#### 4.1.1. Positive perceptions of overlay designs

Four primary themes were identified in participants’ explanations for their preferred overlay design: unobtrusiveness; clarity and structure boundary recognition; sufficiency of information; and safety and risk awareness.

- **Unobtrusiveness:** Outline was selected as the most preferred design by twenty participants for the sella and by eighteen for the parasellar structures. A primary reason participants favoured the outline design was its minimal obstruction of the surgical field. Most participants appreciated the Outline design for its ability to clearly delineate anatomical boundaries without obscuring the surgical field. It was described as a minimal overlay that maintained anatomical visibility and reduced visual distraction by avoiding unnecessary clutter or masking critical areas. For example, P8 described it simply as *“it does not obstruct the anatomy”*, and P34 noted that it provided *“all the information with the least overlay of normal structures”*. Several participants also mentioned that the Outline felt less tiring to the eye and helped preserve focus during the task, including P17, who noted that it gave *“clear information without being tiring to the eye and without covering the normal anatomical structures with colour”*.
- **Clarity and structure boundary recognition:** A primary reason for participants’ design preference was the ability to convey anatomical boundaries and facilitate structure recognition. This was most frequently associated with the Outline design for both the sella and parasellar regions. Eleven participants specifically noted that it helped define clear margins and distinguish structures without obscuring the underlying anatomy. For example, P21 noted that it *“gives a good indication of the structure without obstructing the view”*, and P30 commented that it *“showed the boundaries, but does not mask the structure”*. Others said the outline helped define the working field (P15), estimate the extent of bone removal (P38), or identify specific landmarks like the carotid arteries (P25). For some, it provided a general overview of the anatomy without clutter, described as *“synoptic”* (P5) or simply *“clear”* (P24). Some participants also valued the Mask for conveying the full anatomical volume in a simple, unobtrusive way. P18 described the mask as offering *“a nice visualisation of the whole structure”*, and P16 mentioned that it supported a better sense of 3D orientation. A few participants also noted the clarity of the mask, with P28 finding the colour-coded overlay easy to interpret. Although mentioned less frequently, the Heatmap was seen by some as useful for spatial understanding. P18 noted that it provided *“a nice picture of where everything is and the relationship to the sella”*.
- **Sufficiency of information:** A key factor in participants’ preference for certain sella or parasellar designs was the sense that they provided sufficient information. Seven participants who selected the Outline design across both regions noted that it displayed just enough detail to understand the anatomy without overwhelming the view. P34 noted that it offered *“all necessary information with the least overlay of the normal view”*, while P23 described it as *“clear and adds confidence”*. Others highlighted that it helped them visualise danger zones and adjust their approach accordingly. Three participants mentioned the Heatmap in relation to the amount of information it conveyed. P37 felt it was more informative than other options, as it *“adds a degree of confidence”*, whilst P28 and P29 saw the level of detail and uncertainty gradient as helpful for training, especially for younger or less experienced surgeons.
- **Safety and risk awareness:** Participants who preferred the Heatmap and Mask designs emphasised their effectiveness in communicating risk levels and enhancing safety. Eleven participants noted that these designs helped identify danger zones and avoid critical structures, with the colour scheme aiding in the recognition of areas requiring caution. P38 noted that the colour made it easier to *“recognise areas at risk”*, whilst P6 appreciated that it helped with *“ensuring safety during the procedure”*. Others commented on the value of gradual or graded markings in the Heatmap, with P9 describing it as showing a *“gradual border between target structure and risky structures”*, and P23 noting that it *“reflects a degree of uncertainty of sella location”*. P14 added that seeing these danger zones helped them *“think about approach a bit more and wider considerations”*. A few also mentioned that these designs helped reinforce which areas are usually safe to enter, especially when working close to high-risk regions.

#### 4.1.2. Negative perceptions of overlay designs

The negative feedback provided by participants highlighted two broad themes that explain why certain designs were less preferred: lack of clarity or relevance; and visual obstruction and noise.

- **Lack of clarity or relevance:** A common reason for disliking certain designs was their perceived lack of clarity, relevance, or usefulness to the task. Several participants noted that some overlays, particularly the centroid design, failed to provide additional valuable information and merely repeated what was already known. P18 said the centroid was *“the least informative and presented information I already knew and therefore it is more noise than helpful”*, while P37 noted that it *“can be matched by anatomical knowledge or neuronavigation alone”*. Others commented on a lack of clear structure boundaries. For example, P6 stated that their least preferred design *“does not delineate the at risk structure”*, and P31 described it as one that *“does not define limits”* and *“does not change what we already know”*. Some were also criticised for lacking operative usefulness. P12 referred to one option as *“just a label, not very helpful operatively”*, while P35 felt it was *“not really operatively relevant, doesn’t add much to the safety or approach”*.
- **Visual obstruction and noise:** Twelve participants presented concerns related to visual obstruction or distraction. Several mentioned that Heatmap and Mask designs interfered with their view of the underlying anatomy. P8 said it *“distracts from the surgical anatomy”* while P24 explained that it *“obscured the view”* and *“masked the anatomy”*. Others pointed to the amount of colour or shading as a source of visual clutter. P13 described their least preferred option as one that *“obstructs the actual anatomy due to too much shading”*, and P15 felt that for the heatmap *“I don’t know what the different colours represent.”* and *“The different colours would be too distracting and potentially confusing”*. For some, this made the information harder to interpret. P17 described the textual descriptions as *“confusing to read”* and P23 said that *“colours were similar”* and made the design feel *“very messy”*. P4 summarised the tension clearly by noting that *“too much info kills the info”*.

#### 4.1.3. Suggested Improvements

Four main themes emerged from participants’ suggestions for improving overlay designs:

- **Improvement of visual style and colour properties:** Fourteen participants suggested improving colour contrast (P11, P27, P36), using more defined or relevant colour schemes for different structures (P4, P21, P32), and allowing for customisation options such as toggling colours or adjusting opacity (P5, P12). Several participants requested clarification of colour meaning, particularly for critical structures, and others highlighted the need for more consistent and accessible colour design. As one participant suggested, *“Maybe [add the] ability to change colour, solid versus dotted line etc”* (P36). Across responses, there was a consistent request for greater visual precision and flexibility, with improved resolution (P21), clearer boundaries (P28), and the ability to adapt visual elements to the surgical context (P36).
- **Incorporate flexible design modes:** Twelve participants suggested improvements focused on combining or toggling between different designs. Rather than relying on a single mode, integrating multiple designs may improve clarity and contextual understanding. Suggestions included combining the clear structural boundaries of the Outline with the shading or gradual transitions offered by Heatmap or Mask (P4, P21, P22). Others emphasised the value of switching designs on demand depending on the task, either manually or through input controls. As one participant noted *“Toggle off and on or toggle between Heatmap and Outline”* (P37) would enhance the system’s flexibility. This desire for flexible switching was echoed by others who proposed *“cycling between Mask/Outline/Off “* (P11) or temporarily activating overlays such as the Heatmaps when needed (P34).
- **Enhancing perceived accuracy and safety:** Fourteen participants provided suggestions aimed at improving how the designs reflect anatomical accuracy and support safe decision-making. Some focused on increasing precision; for example, by making outlines of high-risk structures more accurate or clearly marked (P12, P35). A few participants mentioned the potential to indicate margins of confidence, such as using multiple boundary lines to show where the system is more or less certain. As one participant explained, *“Marking of margin of confidence/margin of error, where is the computer more confident and where is it less confident?”* (P15). The need for better depth perception was also present in their comments, with suggestions to incorporate 3D views to help visualise complex structures like the optic nerves or parasellar carotids (P17, P18, P32). In two cases, participants expressed a desire to cross-check the visualisations with real-time data sources *“I would be more satisfied if this system could be merged in real-time with Doppler ultrasound during surgery to increase its reliability”* (P7), or to integrate it with a navigation system (P26).
- **Feedback and adaptivity:** Three participants made suggestions related to how the visualisations could respond or adapt in real time. For example, P2 suggested integrating real-time updates from other imaging modalities to improve accuracy, while P13 asked for the Outline to remain dynamic as the view changes, with borders adjusting to shifting perspectives. P7 described a desire for the system to provide reassurance *“showing that I am on the right track”* indicating a need not only for visual guidance but also for ongoing confirmation during navigation.

### 4.2. Overlay Triggers

Regarding the preference for overlay triggers, the user-enabled trigger was ranked highest on average and most frequently ranked first, while the always-on trigger was ranked lowest on average and most frequently ranked last (Table 3). Subgroup analysis showed no statistically significant difference between subgroups with Friedman test p=0.999 for each group; Mann-Whitney U test: p=1.0 and OLR effect of group on ranking p=0.903. Wilcoxon Signed Rank testing revealed no statistically significant preference for user-enabled (sella & parasellar structures) trigger over the second highest ranked instrument-triggered (parasellar) & sella user-enabled with p=0.482. Always-on (sella & parasellar structures) trigger was the least preferred with statistical significance over all other triggers.

**Table 3:** Analysis of overlay trigger rankings; Rows are ordered by quartile statistics; User-enabled (sella & parasellar) was the highest ranked, though not significantly different from instrument-triggered (parasellar) & sella user-enabled (bold text); Always on (sella & parasellar structures) is significantly the least preferred over others (underlined). PU-SU: user-enabled (sella & parasellar structures); PI-SU: instrument-triggered (parasellar) & sella user-enabled; PI-SA: instrument-triggered (parasellar) & sella always on; PA-SA: always-on (sella & parasellar structures)

| Trigger | Count |  |  |  | Quartile statistics |  |  | Wilcoxon test - p value |  |  |  |
| --- | --- | --- | --- | --- | --- | --- | --- | --- | --- | --- | --- |
|  | Rank 1 | Rank 2 | Rank 3 | Rank 4 | Median | Q1 | Q3 | PU-SU | PI-SU | PI-SA | PA-SA |
| PU-SU | 19 | 6 | 10 | 3 | 1.5 | 1 | 3 | - | **0.482** | 0.015 | 0.000 |
| PI-SU | 12 | 15 | 4 | 7 | 2 | 1 | 3 | **0.482** | - | 0.053 | 0.001 |
| PI-SA | 4 | 10 | 19 | 5 | 3 | 2 | 3 | 0.015 | 0.053 | - | <u>0.013</u> |
| PA-SA | 3 | 7 | 5 | 23 | 4 | 2.25 | 4 | 0.000 | 0.001 | <u>0.013</u> | - |

#### 4.2.1. Helpfulness vs distraction

Table 4 presents the quantitative analysis of participants’ rating each trigger on helpfulness and distractibility in a 5-step Likert scale. This comparison revealed that the User-enabled trigger was perceived as the most helpful, with statistical significance over instrument-triggered (parasellar) & sella always on (p=0.039). In contrast, the least helpful was instrument-triggered (parasellar) & sella user-enabled, which was rated lower than always-on (sella & parasellar structures) with statistical significance (p<0.001). The most distracting trigger was instrument-triggered (parasellar) & sella user-enabled, significantly more so than always-on (sella & parasellar structures) with p<0.001. Conversely, user-enabled (sella & parasellar structures), was the least distracting over instrument-triggered (parasellar) & sella always on with statistical significance (p=0.002). The inverse correlation between helpfulness and distractibility rankings, supported by statistical analysis, aligns with expectations. The most helpful and the least distracting trigger, user-enabled (sella & parasellar structures) also aligns with the quartile statistics in Table 3, reaffirming user-enabled (sella & parasellar structures) as the highest ranking trigger in the absence of statistical significance.

**Table 4:** Analysis of Helpfulness vs Distractibility of overlay triggers; Rows are ordered by mean (descending for helpfulness and ascending for distractibility); User-enabled (sella & parasella structures) is the most helpful and least distracting trigger with statistical significance over others (bold text); PU-SU: user-enabled (sella & parasellar structures); PI-SU: instrument-triggered (parasellar) & sella user-enabled; PI-SA: instrument-triggered (parasellar) & sella always on; PA-SA: always-on (sella & parasellar structures)

| Trigger | Helpfulness |  |  |  |  | Distractibility |  |  |  |  |
| --- | --- | --- | --- | --- | --- | --- | --- | --- | --- | --- |
|  | Mean | Wilcoxon test - p value |  |  |  | Mean | Wilcoxon test - p value |  |  |  |
|  |  | PU-SU | PI-SA | PA-SA | PI-SU |  | PU-SU | PI-SA | PA-SA | PI-SU |
| PU-SU | 4.367 | – | **0.039** | 0.003 | 0.000 | 1.868 | – | <u>0.002</u> | 0.001 | 0.000 |
| PI-SA | 3.921 | **0.039** | – | 0.567 | 0.000 | 2.632 | <u>0.002</u> | – | 0.549 | 0.000 |
| PA-SA | 3.763 | 0.003 | 0.567 | – | <u>0.000</u> | 2.711 | 0.001 | 0.549 | – | **0.000** |
| PI-SU | 2.342 | 0.000 | 0.000 | <u>0.000</u> | – | 4.211 | 0.000 | 0.000 | **0.000** | – |

#### 4.2.2. Positive perceptions of overlay triggers

Participant responses revealed three key positive themes across their preferred visualisation trigger types. Their preferred triggers provided either some or all of the following: visual clarity and cognitive ease; user control and flexibility; and support for safety and confidence.

- **User control and flexibility:** A key theme was that participants felt their preferred trigger provided a sense of control or agency, with 20 participants emphasising this aspect. User-enabled (sella & parasellar structures) (14 participants) achieved the highest ranking and was identified as the most helpful and least distracting. As described by P2: *“I liked the user-enabled trigger because it offers full control, allowing activation only when needed, which minimises distractions and ensures the visualization appears precisely when it’s most relevant during the procedure”*. Five participants who selected instrument-triggered (parasellar) & sella user-enabled also pointed out the capability of this one to allow them to be in control while providing some assistance: *“The other structures enabled with the instrument reminds you when you stray, but do not distract you”* (P13). In addition, two participants referred to user-enabled (sella & parasellar structures) as versatile and flexible, while P28 said, for instrument-triggered (parasellar) & sella user-enabled trigger, *“Choice allowed for the sella assessment based on anatomy, and would allow for human judgement in cases of anatomical variation”*.
- **Visual Clarity and cognitive ease:** Several participants appreciated triggers that reduced distraction and supported focus. Instrument-triggered (parasellar) & sella user-enabled was most often described as less distracting (5 participants) and less obtrusive (2 participants), helping maintain clarity by appearing only when relevant. One participant described this as *“balance between assistance and distraction”*(P27) while another said it *“didn’t distract until required”* (P24). User-enabled (sella & parasellar structures) was also seen as cognitively easy to manage, with 2 participants finding them less distracting, two calling them less obtrusive, and one noting their consistency. P2 explained this trigger as it *“ […]minimizes distractions and ensures the visualization appears precisely when it’s most relevant”*.
- **Support for safety and confidence:** Several participants valued triggers that offered balanced assistance by providing timely, context-aware support without taking over the procedure. The trigger, instrument-triggered (parasellar) & sella user-enabled, in particular, was seen as helpful for enhancing safety while allowing the surgeon to remain in control. As P33 noted, it *“gives independence, not too leading, but still supportive, not distracting, lets you think”*. This visualisation also increased confidence by offering reassurance at critical moments, such as when deciding whether to remove more bone or when drifting off track (P17, P25). P37 described a similar sense of confidence when using user-enabled overlays, as it allowed them to *“remove visualisations when I was comfortable and then toggle the sella in as a guide when I was unsure, choosing to keep parasellar structures permanently visualised”*. In addition, two other participants noted that user-enabled (sella & parasellar structures) trigger provided an increased feeling of safety (P38 and P1).

#### 4.2.3. Negative perceptions of overlay triggers

Participants’ justifications for their least preferred overlay designs highlighted the following themes.

- **Distraction and visual clutter:** The most common reason participants disliked their least preferred option was its distractibility, particularly among those who selected the always-on (sella & parasellar structures) trigger as their least preferred. Fifteen participants expressed concerns about this, with eight participants specifically mentioning that the constant presence of the overlay was distracting, especially when the information was unnecessary. Some also mentioned that it could obstruct the view of the real anatomy during surgery and provoke annoyance (P13), or even overwhelm them. As P17 put it: *“too crowded to the eye with lots of information that are not needed at all times”*. Three participants expressed similar concerns about distraction in relation to the instrument triggers. P2 explained: *“I disliked the instrument-triggered (parasellar) & sella user-enabled triggers because they can activate unintentionally during movements, leading to distractions and interruptions in focus”*.
- **Lack of user control:** Users disliked the triggers that diminished their sense of agency. This was the most common criticism (four participants) expressed by those who chose always-on (sella & parasellar structures) as their least preferred, with all of them commenting they would rather choose when to display the AR. The lack of user control was also noted by two participants who disliked Instrument triggers, for which P2 explained: *“they lack the flexibility of precise timing, as they rely on pre-set criteria rather than real-time decision-making by the user”*.
- **Disrupted system and decision making confidence:** Some participants remarked that the instrument-triggered and always-on triggers had an impact on their decision making confidence. For P15, always-on (sella & parasellar structures) could induce second-guessing: *“if confidently drilling beyond the suggested outline induces self-doubt and second-guessing to an unacceptable degree”*. P33 and P36 expressed the view that the always-on (sella & parasellar structures) could induce a false sense of safety or be *“too leading, [you] lose the opportunity to try yourself “* (P33). Additionally, for P7 and P37, the instrument-triggered visualisations reduced confidence in the system, given that the triggered information is not visible at all the times when it may be needed (e.g., when close to the structure but just out of the trigger threshold).

#### 4.2.4. Suggested improvements

Several participants suggested potential improvements for overlay triggers. Four participants suggested the use of voice activation instead of a toggle button to display the overlay, with P21 mentioning: *“I would improve the user-enabled trigger by adding customisable gesture or voice commands for quicker, hands-free activation, allowing seamless integration into the workflow without needing physical interaction”*. Five other participants suggested including an additional source of feedback (visual, haptic or audio) as an improvement or in place of the instrument-triggered. P38 would like the visualisation to flash when approaching the parasellar structures, and P30 suggests *“it would be even better if the mask disappears as the dura is exposed. i.e. masks reflect the drilling”*. Among those who prefer additional feedback, P7 and P12 perceive it as a warning sound or voice indicating proximity to critical structures; P38 suggests a *“parking sensor type noise”*, while P1 envisions it as an invisible wall (haptic feedback), describing it as *“something that stops the drill when it is close to a critical structure”*.

### 4.3. Overlay Placement

The overlay below the instrument (underlay) was preferred by the majority of participants (63.2%, 24/38), compared to 31.6% (12/38) of participants who preferred overlay placement over the instrument. Two participants did not have a preference. Post-hoc analysis via Wilcoxon Signed Rank testing did not reveal a statistically significant preference for overlay placement. The subgroup analysis did not show a statistically significant difference between subgroups (Friedman test p=1.0 for each group; Mann-Whitney U test: p=1.0 and OLR effect of group on ranking p=1.0). However, while residents/fellows did not have a statistically significant preference (9 and 8 for underlay and overlay with a p=0.854), consultants preferred underlay with p=0.026.

Regarding the open-ended feedback, participants provided several reasons for preferring one placement over the other. For those who favoured the overlay over the instrument, two main reasons emerged. Six participants stated that it made the AR clearer and more visible, with P14 noting the additional benefit that an overlay *“[…] may not be obscured during surgery if blood/wash/etc. used”*. Three participants stated that this visualisation is more logical or natural. From those who did not prefer overlay over the instrument, P17 stated *“I think that AR visualization over the instrument is distracting and it gives the impression that some damage has already been done”*. In contrast, the majority of participants preferred the overlay below the instrument. A few clear themes were identified in their responses. The most common theme (8 participants) was that it provides better visibility of the instrument, with P18 explaining: *“I can see the outline of what I’m drilling”*. Another salient theme was that the underlay was more in tune with real-life expectations of the AR (7 participants), with participants describing it as more natural, or realistic. For P11, *“it makes more sense, otherwise a bit strange optically”*. Additionally, four participants expressed that the underlay minimised obstruction and distraction. Another four participants expressed that underlay conveyed a sense of safety and control: *“[…] it gives an impression of safety and protection (if that makes sense) like a layer between the instrument and the important structures”* (P17).

### 4.4. Other Reflections and Additional Feedback

A small number of participants changed their answers to preferred overlay designs after performing the interactive tasks for testing overlay triggers. For the sellar region, two participants changed their preference: one from Outline to Mask, and another from Heatmap to Outline. For the parasellar structures, two participants changed their preference from Centroid to Mask. However, these participants did not justify this change of choice. In addition, while P7, P14 and P34 did not change their preference after interacting with the overlay triggers, they did point out that their choices may change when testing in real life. Several participants expressed positive impressions of the different overlay designs. Comments described the system as *“very useful”* (P8, P9), with others praising the concept and design quality more explicitly. P13 noted, ‘*‘The designs are great, I would totally give them a try in the OR”*, while P17 described the study as *“brilliant”* and *“a great test of this exciting tech”*. Participants also acknowledged progress over previous iterations (P18) and highlighted the potential clinical value, particularly concerning safety and real-world surgical integration. As P38 noted, *“It’s an excellent idea. Anything to improve patient safety is a great idea”*.

## 5. Discussion

This study engaged an international cohort of surgeons, assessing their preferences for the design of AR systems to support endoscopic surgery, focusing on three key aspects of visualisation: overlay design, overlay trigger and overlay placement relative to the surgical instrument. The visualisations used in this study were applied to two types of regions: the sella, a central bony landmark commonly targeted for resection, and the parasellar structures, which represent surrounding anatomy to be preserved. This framing enabled us to capture preferences for visualising both areas to operate on and areas to avoid. While previous studies have demonstrated potential benefits of AR in neurosurgery, there has been limited research into how these visual elements should be designed to align with real surgical workflows. By capturing quantitative rankings alongside qualitative feedback, this study contributes practical, user-grounded insights for AR systems that support clarity and safety in complex surgical environments.

### 5.1. Overlay Design: Balancing Clarity and Spatial Information

Surgeons demonstrated a clear preference for Outline overlays, particularly for central structures like the sella, where designs should be salient but not distracting. Participants valued outlines for their ability to highlight structural boundaries without obstructing the surgical view. In contrast, Centroid overlays were consistently ranked lowest and described as too abstract or sparse to offer meaningful guidance. These designs were included based on prior work demonstrating their potential as a computationally efficient baseline [11], but most participants did not find them informative enough in practice.

For more complex regions such as the parasellar area, feedback indicated that additional spatial cues were valued. Some participants preferred Mask or Heatmap overlays in this context, noting that these offered a better sense of anatomical volume and “danger zones”. However, Heatmaps were also criticised for contributing to visual clutter, especially when applied across larger structures. These comments reflect a broader challenge in AR interface design: how to provide helpful spatial information without overwhelming the user. This tension is consistent with literature on cognitive ergonomics and interface clutter in high-stakes settings [3, 34, 35].

Participants also requested greater flexibility in adjusting the visualisation style. Features such as the ability to toggle between overlay types, modify line thickness or adjust colour and opacity were frequently mentioned. These suggestions support calls in the surgical HCI literature for modular, user-configurable AR interfaces [36, 19].

### 5.2. Overlay Triggers: Supporting Autonomy Without Disruption

Triggers should allow seamless user-control. There was strong consensus in favour of user-controlled triggers, which were seen as the least distracting and most intuitive option. Surgeons appreciated the ability to activate overlays on demand, which allowed them to maintain focus and workflow rhythm. Always-on overlays, although more predictable, were sometimes described as tiring or mentally taxing over longer periods.

Instrument-triggered overlays received mixed reactions. While some participants found the automation appealing, others felt it introduced unnecessary distractions or behaved unpredictably, especially when applied to sella overlays. These findings echo previous concerns in the literature about automation reducing user confidence in the system or creating mismatches between system logic and surgeon intent [37, 38].

Participants emphasised the importance of control: when AR systems behave in ways the user does not anticipate, even if technically correct, they may be perceived as less trustworthy. Some suggested hybrid approaches, such as optional prompts or “nudges” that appear in context but can be dismissed. Voice activation was also mentioned as a possible hands-free interaction method, building on findings from surgical studies that explored gesture and voice-based control [23].

### 5.3. Overlay Placement: Navigating Visibility and Depth

Preferences for overlay placement were more variable, but placing overlays beneath the surgical instrument was most frequently ranked first. This configuration was perceived as aligning better with intuitive depth cues and preserving visibility of the tool. However, participants who preferred placing the overlay on top of the instrument described specific intraoperative challenges, such as blood wash, fogging, or lighting artefacts, that obscured anatomical landmarks. In such cases, top-layer overlays helped retain spatial reference points.

These findings suggest that overlay placement should be adaptable, allowing users to switch between placement modes depending on intraoperative conditions. Whether this switching should be manual or automatically triggered (e.g., via detection of visual occlusion) remains an open question. Nonetheless, the emphasis on flexibility reflects findings in related domains, where surgeons prefer AR systems that respond to variability in surgical conditions [39, 40].

### 5.4. Broader Implications for AR Design in Surgery

Across all domains, participants voiced a clear preference for systems that support accurate anatomical navigation, minimise distraction, and allow for individual control of information flow. These themes mirror established HCI and human factors principles [37, 34], and have been echoed in the AR surgery literature, particularly in studies of spine and laparoscopic procedures [36, 41, 42].

Importantly, several participants described AR as a complement, not a replacement, for expert decision-making. They stressed the value of overlays that reinforce confidence rather than introduce doubt. This aligns with emerging models of surgical technology adoption, which advocate for interfaces that act as cognitive aids rather than autonomous decision-makers [43, 38].

Participants also shared forward-looking expectations. Some suggested that overlays should be dynamically updated in real time and integrated with adjunct technologies such as Doppler ultrasound or neuronavigation systems. These comments align with broader technological trends toward “smart” operating rooms and AI-supported guidance tools [43, 44].

Notably, preferences appeared consistent across levels of surgical experience. This suggests that the findings reflect widely shared needs rather than biases tied to seniority or familiarity with technology. As AR systems advance, these user-centred insights provide a foundation for designing tools that support safe, usable, and customisable surgical guidance.

### 5.5. Strengths and Limitations

As surgeons with expertise in the eTSA and pituitary adenoma resection represent a small, time-constrained population, we developed an online survey with interactive features to achieve a balance between accessibility and data richness. Its international reach enabled engagement with a niche group of specialists across multiple regions, and the interactive design helped participant engagement. Despite our efforts in international survey dissemination, the majority of respondents were from the UK and high income countries, representing the difficulty of accessing these busy professionals who have limited free time, especially in resource constrained settings. While all respondents completed the demographic, ranking, and Likert-scale questions, approximately 75% of open-ended responses across the survey were considered substantive, with the remainder consisting of minimal entries such as “N/A” or single-word answers. The median number of non-substantive responses per question was 5, indicating some variability in engagement across the 11 open-ended items. This highlights a key limitation of survey-based methods, which lack the opportunity for real-time dialogue and probing found in interviews. Nevertheless, in-person studies would likely have been limited to clinicians from a single centre, whereas this approach allowed for broader input. Furthermore, our qualitative analysis was limited to one coder and therefore no measures of agreement were used.

As most centres perform these procedures with 2D endoscopes and there are currently no accurate 3D-2D registration techniques used in clinical practice which allow accurate and automatic 3D overlay onto live 2D surgical video, our virtual simulation was restricted to 2D overlays on static 2D images. As described in the results section, some participants expressed uncertainty about their preferences, particularly regarding overlay triggers and the placement of overlays in relation to the instrument, until they had the opportunity to explore the system further. Therefore, while our findings are informative, they may not fully translate to live surgical environments involving real-time video feeds, where surgeons derive 3D understanding of the boundaries of critical structures from visual cues (e.g. contours and shadows generated by the endoscope’s light), and proprioceptive and haptic feedback from instruments. Further validation is needed through physical simulations and real-time surgical contexts, specifically to assess the impact of these AR design changes on surgical safety and performance, and whether changes in preferences manifest during real-world deployment, particularly during adverse events or atypical cases.

## 6. Conclusion

This study is the first structured evaluation of surgeon preferences for AR overlay design, trigger mechanisms, and placement strategies using eTSA as an example of a high-stakes surgery. By combining quantitative rankings with qualitative thematic analysis, we identified clear user priorities: overlays should maintain clarity without obstruction, trigger activation should remain under user control to avoid cognitive disruption, and placement should align with natural depth perception while allowing customisation for changing intraoperative conditions. These findings highlight that effective surgical AR must prioritise usability and be aligned with surgical workflow. Future development should focus on validating these design principles in higher-fidelity real-time simulations and live surgical environments, with the goal of advancing AR systems that support safer, more efficient surgical practice.

## Data Availability

Available upon request

## Acknowledgements

This work received funding from the UCL Hawkes Institute - formerly the Wellcome/EPSRC Centre for Interventional and Surgical Sciences (WEISS) (203145/Z/16/Z)-, the EPSRC grant CARES: Context aware Augmented Reality for Endoscopic transsphenoidal Surgery (EP/W00805X/1), the EPSRC grant AID-PitSurg: AI-enabled Decision support in Pituitary Surgery (EP/Y01958X/1) and the UCLH/UCL Biomedical Research Centre. We thank members of the CARES project team for their continuous feedback throughout the development of this research. DZK is supported by the Cleveland Clinic Foundation. HJM is supported by the NIHR Biomedical Research Centre at University College London.

## Collaborators

We thank all members of the CARES Evaluation Group for their participation in this study. Collaborative authors are listed in alphabetical order by last name:

- **Agostini, Ludovico** — Fondazione Policlinico Universitario Agostino Gemelli IRCCS, Italy
- **Ali, Ahmad M. S.** — Department of Neurosurgery, The Walton Centre NHS Foundation Trust, Liverpool, UK
- **Al Barajraji, Mejdeddine** — Department of Neurosurgery, Hôpital Citadelle, Liège, Belgium
- **Ayça Şahin, Saime** — University of Health Sciences Turkey, Faculty of Medicine, Istanbul Sisli Hamidiye Etfal Health Training and Research Hospital, Neurosurgery Department, Turkey
- **Bahl, Anuj** — Hull University Teaching Hospitals, UK
- **Baraka, Mohammad** — Cairo University, Egypt
- **Barrit, Sami** — Department of Neurosurgery, Université Libre de Bruxelles, Belgium
- **Borg, Anouk** — University College London Hospital, UK
- **Cearns, Michael D.** — The Walton Centre, Liverpool, UK
- **Chari, Aswin** — Department of Neurosurgery, National Hospital for Neurology & Neurosurgery, UK
- **Decramer, Thomas** — University Hospitals Leuven, Belgium
- **Dow, Graham** — Queens Medical Centre, Nottingham, UK
- **Elserius, Anne** — Royal Stoke University Hospital, UK
- **Giannis, Theofanis** — National Hospital for Neurology and Neurosurgery, UK
- **Hanrahan, John Gerrard** — University College London, Hawkes Institute, UK
- **Jain, Abhiney** — University College London, UK
- **Jalalod’din, Hasan** — Ain Shams University, Cairo, Egypt
- **Jeyaretna, Sanjeeva** — Oxford University Hospitals NHS Foundation Trust and Nuffield Department of Clinical Neurosciences, Oxford University, UK
- **Layard Horsfall, Hugo** — National Hospital for Neurology and Neurosurgery / University College London / Francis Crick Institute, UK
- **Ley, Luis** — Hospital Universitario Ramón y Cajal, Spain
- **Lombard, Arnaud** — CHR Citadelle, Belgium
- **Maghrabi, Yazid** — Neurosciences Department, King Abdulaziz Medical City - Health Affairs, Jeddah, Saudi Arabia
- **Maratos, Eleni** — King’s College Hospital NHS Foundation Trust, UK
- **Martin, Andrew James** — Atkinson Morley Wing, St George’s Hospital, London, UK
- **Mellal, Amine** — Department of Neurosurgery, University Hospital of Lausanne and University of Lausanne, Switzerland
- **Newall, Nicola** — National Hospital for Neurology and Neurosurgery, London, UK
- **Okasha, Mohamed** — Ninewells Hospital and Dundee Medical School, UK
- **Price, Sally-Ann** — Royal Victoria Infirmary, Newcastle, UK
- **Remacle, Thibault** — Department of Neurosurgery, Hôpital de la Citadelle, Liège, Belgium
- **Robins, James MW** — Leeds General Infirmary, UK
- **Sheikh, Asim** — Leeds General Infirmary, UK
- **Sinha, Saurabh** — No affiliation at present
- **Starup-Hansen, Joachim** — University College London, UK
- **Teo, Mario** — Bristol Institute of Clinical Neuroscience, Southmead Hospital, Bristol, UK
- **Tsermoulas, Goergios** — Queen Elizabeth Hospital Birmingham, UK
- **Williams, Adam** — North Bristol NHS Trust, UK

## Appendix A. Survey Questions

### Appendix A.1. Participant Background

In which country are you currently based? __________

Please select from below your level of experience:

- Neurosurgery consultant or attending
- Neurosurgery senior fellow (post-training)
- Neurosurgery registrar or resident
- Other __________

How many years of experience do you have performing the endoscopic transsphenoidal approach? __________

### Appendix A.2. Overlay Display

Please rank the sella visualisations from most to least preferred (drag them to order them).

- _____ Centroid
- _____ Outline
- _____ Mask
- _____ Heatmap

What did you like about your preferred sella visualisation? __________

What would you improve from your preferred sella visualisation? __________

What did you dislike about your least preferred sella visualisation? __________

Please rank the parasellar visualisations from most to least preferred:(drag each item to order them).

- _____ Centroid
- _____ Outline
- _____ Mask
- _____ Heatmap

What did you like about your preferred parasellar display? __________

What would you improve from your preferred parasellar display? __________

What did you dislike about your least preferred parasellar display? __________

### Appendix A.3. Overlay Triggers

I found the always on (sella & parasellar structures) trigger helpful in completing the task.

- Strongly disagree
- Somewhat disagree
- Neither agree nor disagree
- Somewhat agree
- Strongly agree

The always on (sella & parasellar structures) trigger distracted me while completing the task.

- Strongly disagree
- Somewhat disagree
- Neither agree nor disagree
- Somewhat agree
- Strongly agree

I found the user-enabled (sella & parasellar structures) trigger helpful in completing the task.

- Strongly disagree
- Somewhat disagree
- Neither agree nor disagree
- Somewhat agree
- Strongly agree

The user-enabled (sella & parasellar structures) trigger distracted me while completing the task.

- Strongly disagree
- Somewhat disagree
- Neither agree nor disagree
- Somewhat agree
- Strongly agree

I found the instrument-triggered (parasellar) & sella always on trigger helpful in completing the task.

- Strongly disagree
- Somewhat disagree
- Neither agree nor disagree
- Somewhat agree
- Strongly agree

The instrument-triggered (parasellar) & sella always on trigger distracted me while completing the task.

- Strongly disagree
- Somewhat disagree
- Neither agree nor disagree
- Somewhat agree
- Strongly agree

I found the instrument-triggered (parasellar) & sella user-enabled trigger helpful in completing the task.

- Strongly disagree
- Somewhat disagree
- Neither agree nor disagree
- Somewhat agree
- Strongly agree

The instrument-triggered (parasellar) & sella user-enabled trigger distracted me while completing the task.

- Strongly disagree
- Somewhat disagree
- Neither agree nor disagree
- Somewhat agree
- Strongly agree

Please rank the overlay triggers from most to least preferred:

- ___ Always-on (sella & parasellar structures)
- ___ User-enabled (sella & parasellar structures)
- ___ Instrument-triggered (parasellar) & sella always on
- ___ Instrument-triggered (parasellar) & sella user-enabled

What did you like about your preferred overlay trigger? __________

What would you improve from your preferred choice of overlay trigger? __________

What did you dislike about as your least preferred overlay trigger? _________

After having interacted with the overlay triggers previously presented, would you change your original choice of preferred sella display (Centroid, Outline, Mask or Heatmap)?

- Yes
- No

Which sella display would you choose instead as your preferred choice? (optional)

- Centroid
- Outline
- Mask
- Heatmap

Please explain why you would change your original choice of preferred sella display (optional): __________

After having interacted with the visualisation triggers previously presented, would you change your original choice of preferred parasellar structures display (Centroid, Outline, Mask or Heatmap)?

- Yes
- No

Which parasellar structures display would you choose instead as your preferred choice? (optional)

- Centroid
- Outline
- Mask
- Heatmap

Please explain why you would change your original choice of preferred parasellar structures display (optional):__________

### Appendix A.4. Overlay Placement

Which placement option would you prefer during surgery?

- AR display OVER the instrument
- AR display BELOW the instrument

Why would you prefer that option? __________

Do you have any additional comments on all the designs you have seen so far? If there is another display type or trigger that you think would be useful but was not presented, please describe it below: __________

## Footnotes

3 https://framer.com/

4 https://scipy.org/

5 https://www.statsmodels.org

6 https://www.maxqda.com/new-maxqda-24

